# Beyond knowledge: Psychosocial, traditional, and structural determinants of exclusive breastfeeding among young mothers aged 15-29 years in Peri-urban Lusaka, Zambia. A Facility-based cross-sectional study

**DOI:** 10.64898/2026.05.27.26354145

**Authors:** Godwill Silupya, Kalonga Mwiinga, Rosemary N. Likwa, Twaambo Hamoonga

## Abstract

Exclusive breastfeeding (EBF) for the first six months is a critical protective practice, yet determinants beyond knowledge among young mothers in peri-urban sub-Saharan Africa remain insufficiently understood. This facility-based cross-sectional study assessed factors associated with EBF among 413 mothers aged 15-29 attending postnatal services at two public facilities in Lusaka, Zambia (Aug-Oct, 2025). Data from structured interviewer-administered questionnaires covered demographic, socioeconomic, cultural, mental health, peer support, and neonatal care knowledge factors. Logistic regression produced adjusted odds ratios (AOR) with 95% confidence intervals (CI). Although 99.5% reported receiving neonatal care education, 71.6% practiced EBF. Mothers aged 25-29 had lower odds of EBF than those aged 15-19 (AOR = 0.17, 95% CI: 0.03-0.99). Married mothers were more likely to exclusively breastfeed (AOR = 4.83, 95% CI: 1.59-14.65). Separated mothers also showed higher odds (AOR = 13.66, 95% CI: 1.89-98.71), although the wide confidence interval indicates substantial uncertainty and its based on a small subgroup (n=13). Formal employment was positively associated with EBF (AOR = 3.94, 95% CI: 1.12-13.85). Avoidance of specific traditional neonatal practices (AOR = 0.14, 95% CI: 0.04-0.53) and not consulting traditional healers (AOR = 0.06, 95% CI: 0.02-0.18) were also independently associated with EBF. Postnatal anxiety showed a strong inverse association (AOR = 0.14, 95% CI: 0.03-0.76). Parity, income, education, neonatal care awareness, and receipt of health education were not independently associated. These findings suggest that EBF in peri-urban Lusaka is shaped more by social, cultural, and psychological influences than knowledge alone, underscoring the need to integrate mental health screening, culturally sensitive counselling, and family-centred support within postnatal services to improve EBF uptake among young mothers in similar settings.

## Introduction

Neonatal mortality remains a major global health challenge, accounting for nearly half of all under-five deaths worldwide. Although under-five mortality declined from 5.0 million deaths in 1990 to 2.4 million in 2019, 47% of these deaths now occur in the neonatal period, with approximately three-quarters occurring within the first week of life¹. Neonatal mortality has declined more slowly than post-neonatal mortality, reflecting persistent gaps in the quality and continuity of maternal and newborn care. This gap is exemplified by the persistent discordance between maternal knowledge of recommended newborn care practices and the actual adoption of these practices. Globally, an estimated 6,700 newborns die each day, the majority in low- and middle-income countries², highlighting the urgent need to understand factors beyond knowledge that influence neonatal care behaviours.

A substantial proportion of neonatal deaths are preventable through evidence-based interventions delivered across the continuum of care. Skilled birth attendance, early initiation and exclusive breastfeeding (EBF), thermal protection, hygienic cord care, and prompt infection management have demonstrated significant mortality reductions^3,2^. Despite increased facility-based deliveries and access to knowledge across sub-Saharan Africa, improvements in service utilisation have not consistently translated into optimal newborn care practices at household level. Early discharge, limited postnatal counselling reinforcement, and sociocultural influences continue to undermine effective practice adoption^4^.

In Zambia, adolescent and young mothers face disproportionately elevated risks of adverse neonatal outcomes compared to older reproductive-age women. Although the country has made notable progress in reducing neonatal mortality from 27 deaths per 1,000 live births in 2018 to 17 per 1,000 live births in 2024^5,6^ age-related disparities persist. Neonatal mortality remains higher among mothers below 20 years (23 per 1,000 live births) and those aged 20-29 years (19 per 1,000 live births) compared to women aged 30 years and above, with Lusaka Province, peri-urban reporting one of the highest neonatal mortality rates (27 per 1,000 live births) lJ.

Despite nearly universal breastfeeding initiation (98%), sustained exclusive breastfeeding (EBF) for the recommended six months remains suboptimal, reflecting a persistent knowledge-practice gaplJ. Young mothers aged 15-29 years often encounter intersecting structural and psychosocial constraints including economic precarity, limited autonomy, early return to income-generating activities, and sociocultural pressures influencing feeding decisions that may undermine adherence to EBF recommendations. While health education remains central to maternal and child health programming, emerging evidence suggests that knowledge alone is insufficient to drive sustained behavioural change³. Psychosocial factors, social norms, and structural conditions may therefore exert stronger proximal influence on infant feeding practices.

Exclusive breastfeeding during the first six months of life is among the most effective and measurable neonatal survival interventions, associated with reduced infectious morbidity and mortality^3,2^. As such, EBF provides a pragmatic proxy for assessing the quality of neonatal care practices at household level.

While several Zambian studies have examined neonatal care practices, most focus on facility-based maternal knowledge, cord care, or thermal care, with limited attention to the combined influence of socio-demographic, cultural, and psychosocial factors on exclusive breastfeeding (EBF) among adolescent and young mothers^2,3,5^. To date, no studies have specifically investigated how maternal anxiety, peer support, and traditional neonatal practices jointly shape EBF adherence in peri-urban Lusaka, a high-density setting with unique structural and sociocultural challenges. This evidence gap limits the development of contextually tailored interventions that account for behavioural, cultural, and psychosocial determinants of EBF.

This study is conceptually guided by the Health Belief Model (HBM), which posits that health behaviours are influenced by perceived benefits, perceived barriers, cues to action, and modifying psychosocial factors^2,5^. By integrating maternal anxiety, peer support, and traditional practices into the analytical framework, the study moves beyond purely knowledge-based explanations to examine how lived sociocultural and structural contexts influence EBF adherence. Accordingly, the study aimed to assess determinants of EBF among mothers aged 15-29 years in peri-urban Lusaka, Zambia, and to situate the findings within regional and global perspectives in similar settings. The global average of EBF is approximately 48% among infants under six months^2,4^, providing context for interpreting adherence levels relative to international and sub-Saharan benchmarks. Specifically, the study evaluated whether socio-demographic characteristics, traditional neonatal practices, maternal mental health, peer support, and neonatal care knowledge independently predict EBF within this population with the aim of improving interventions within these contexts.

## Materials and methods

### Study design

This was a facility-based cross-sectional study design selected to estimate prevalence and examine associations between EBF and demographic, socioeconomic, cultural, psychosocial, and knowledge-related factors. While this design precludes causal inference, it is appropriate for identifying correlates to inform targeted interventions. The study was conceptually informed by the HBM, guiding selection of explanatory variables a priori.

### Study setting

The study was conducted in Mandevu Constituency, Lusaka District, Zambia. Mandevu is a densely populated peri-urban area with an estimated population of 467,744^7^. The area comprises predominantly low-income and informal settlements characterized by limited infrastructure, sanitation challenges, and a largely informal economy. Of eight health facilities providing postnatal services in the constituency (four public and four private), two public facilities (Chipata Level One Hospital and Ng’ombe Health Centre) were randomly selected for inclusion. These facilities represent primary points of maternal and child health service utilization in the area.

### Study population

The study population comprised mothers aged 15-29 years attending postnatal care (PNC) services at selected health facilities in Mandevu peri-urban during the data collection period. This age group was deliberately targeted due to its documented vulnerability to suboptimal neonatal care practices and its demographic contribution to neonatal morbidity and mortality patterns in Zambia^5^. Facility-based recruitment ensured access to mothers within the immediate postnatal window, thereby enhancing the accuracy of self-reported neonatal care practices while maintaining alignment with the study’s cross-sectional design.

Eligible participants were mothers aged 15-29 years who resided in Mandevu, had recently delivered a live birth, were attending PNC services during the study period, were physically and mentally stable at the time of interview, and provided informed consent (with assent and guardian consent obtained for minors). Mothers were excluded if they were younger than 15 or older than 29 years, critically ill or medically unstable, declined or withdrew consent, or were unable to complete the questionnaire due to severe cognitive or communication limitations. These criteria were applied to ensure ethical participation, minimize response bias, and maintain internal validity of the study findings.

### Sample size determination

Sample size was calculated using Cochran’s formula for estimating proportions in cross-sectional studies^8^:

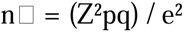

Due to absence of prior facility-based EBF data for this specific subpopulation a conservative prevalence (p = 0.50) was considered with 95% confidence level (Z = 1.96), and 5% margin of error (e = 0.05) to give us the initial sample size calculated at 384. To account for 10% non-response, the target sample was increased to 428 participants.

A total of 413 mothers completed interviews, yielding a response rate of 96%.

### Sampling procedure

A multi-stage sampling approach was employed:

1. **Facility selection:** Two facilities were selected through simple random sampling from eight eligible facilities.
2. **Allocation:** Sample size was proportionally allocated based on average PNC attendance at each facility.
3. **Participant selection:** Systematic sampling was applied within facilities.

At Chipata Level One Hospital approximately 300 eligible mothers where targeted and the sampling interval was approximately 1, resulting in near-consecutive recruitment and selection bias. At Ng’ombe Health Centre approximately 190 eligible mothers where targeted with the sampling interval of 2, resulting in selection of every second eligible mother. A random starting point was selected daily. Replacement was conducted only when selected participants were ineligible or declined participation. The total number of participants interviewed was 413 out of the 428 determined sample size (See figure 1).

**Figure 1:**
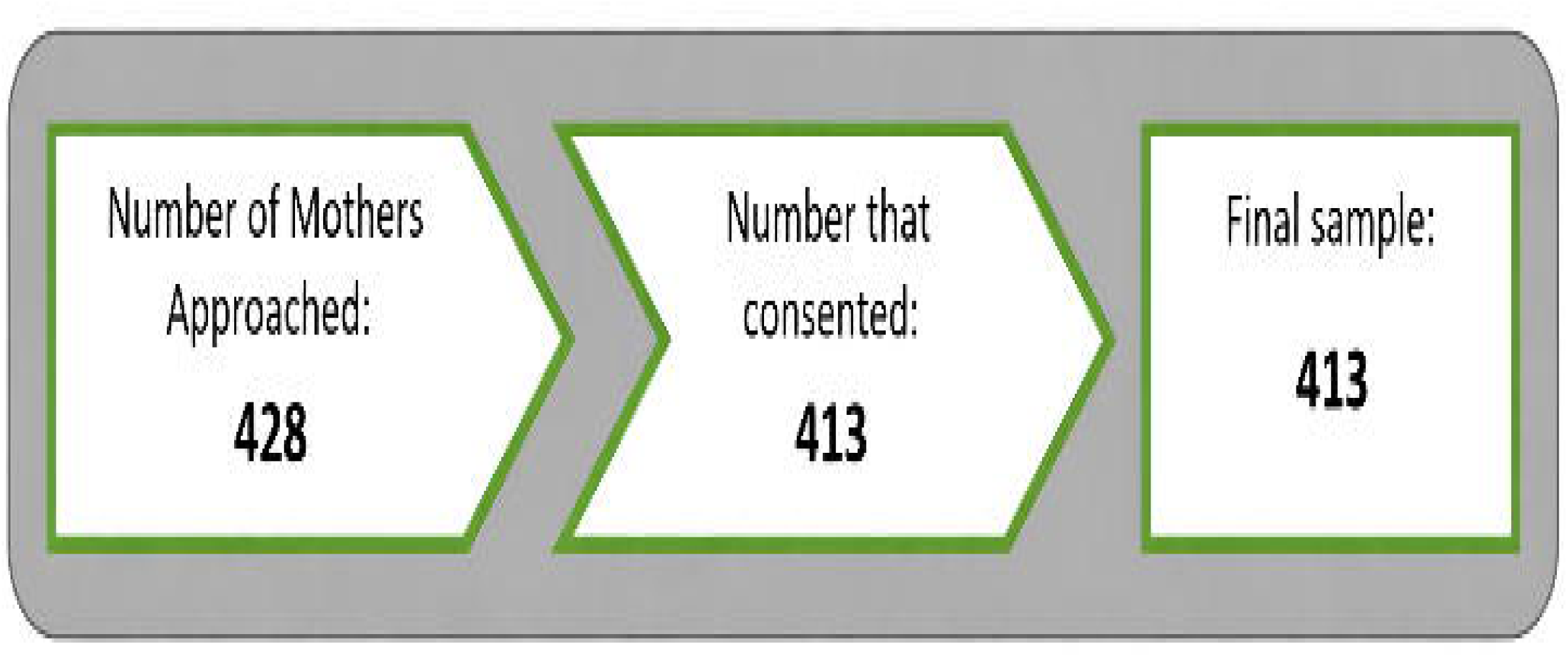
Participant enrolment flow chart.

### Data collection

Primary data were collected using a structured, interviewer-administered questionnaire developed based on neonatal care guidelines and relevant literature, prepared in English, during the period 11^th^ August to 30^th^ October, 2025. The instrument included closed and semi-closed questions covering: i) social-demographic characteristics, ii) social-economic status, iii) traditional neonatal practices, iv) maternal postnatal anxiety, v) peer support, vi) knowledge of neonatal care practices and vii) exclusive breastfeeding practice.

The questionnaire was translated into Nyanja and Bemba and pretested in a comparable peri-urban facility (Kalingalinga Urban Health Centre) to ensure clarity and contextual appropriateness. Necessary revisions were made following pretesting. Back-translation into English was performed by independent bilingual translators to ensure semantic equivalence. Interviews were conducted in participants’ preferred language.

Interviews were conducted face-to-face in private settings to minimise response bias, lasted approximately 15-20 minutes, and completed questionnaires were reviewed daily to ensure completeness and data quality.

### Variables

#### Outcome variable

The primary outcome variable was EBF, defined as feeding the infant only breast milk without the addition of water, other liquids, or solid foods, in accordance with World Health Organization recommendations^2^. EBF was operationalized as a binary variable (yes/no) based on maternal self-report, consistent with standard population-based measurement approaches.

Exclusive breastfeeding (EBF) was assessed using maternal recall since birth rather than the WHO-recommended 24-hour recall method. This approach was adopted to better capture sustained feeding practices over time, as the 24-hour recall method may overestimate EBF prevalence by reflecting only recent behaviour and not continuous adherence since birth. Evidence suggests that point prevalence measures can misclassify infants who were previously given other foods or liquids as exclusively breastfed^9,10^.

However, this method may be subject to recall bias and limits direct comparability with studies using the standard WHO indicator^11^. This limitation is acknowledged, and findings should be interpreted with caution when comparing EBF prevalence across studies using different measurement approaches.

#### Independent variables

Independent variables were categorized into demographic (age, marital status), socioeconomic (education level, household income, employment status), behavioural and cultural (traditional neonatal practices), psychosocial (maternal postnatal anxiety, peer support), and cognitive (knowledge of neonatal care practices) domains. Variable inclusion in the multivariable model was determined a priori based on theoretical plausibility and empirical evidence from existing literature, rather than solely on statistical significance at the bivariate level, to ensure appropriate control for potential confounding and strengthen analytical rigor.

Maternal anxiety was measured using a brief self-reported item rather than a validated psychometric scale; therefore, the measure may not fully capture the clinical complexity of postnatal anxiety and limits direct comparability with studies using standardized instruments.

Internal consistency of the composite knowledge scale was also assessed using Cronbach’s alpha and demonstrated acceptable reliability (α = 0.78).

### Data analysis

Data were analysed using STATA version 17, with statistical significance set at p < 0.05 and 95% confidence interval. Descriptive statistics summarized participant characteristics and the prevalence of EBF, with categorical variables presented as frequencies and percentages.

Unadjusted logistic regression models were fitted to estimate crude associations between each independent variable and EBF. Multivariable logistic regression models were fitted to estimate adjusted odds ratios (AOR) with 95% confidence intervals using an a priori theoretical framework to minimize data-driven variable selection bias and ensure appropriate control of potential confounders. All variables considered conceptually relevant were retained irrespective of univariate significance, consistent with confounder-control principles.

Model fit was assessed using the Hosmer-Lemeshow goodness-of-fit test. Multicollinearity diagnostics were evaluated using variance inflation factors (VIF), with all predictors demonstrating VIF < 10, indicating acceptable independence among explanatory variables.

Several methodological strategies were implemented to minimize bias and strengthen internal validity. Systematic sampling within facilities reduced selection bias, private interviewer-administered questionnaires mitigated social desirability bias, and pretesting enhanced measurement clarity and contextual validity. A priori specification of explanatory variables limited data-driven model overfitting.

Missing data were assessed prior to analysis. As the proportion of missing observations across all variables was below 5%, complete case analysis was performed. Sensitivity assessment indicated no systematic pattern of missingness.

Nonetheless, the facility-based cross-sectional design may limit generalizability to mothers not attending postnatal care services, and reliance on self-reported EBF introduces potential recall and reporting bias.

This cross-sectional study is reported in accordance with the STROBE (Strengthening the Reporting of Observational Studies in Epidemiology) guidelines for observational research. A completed STROBE checklist is provided as Supporting Information (S1).

### Ethical considerations

Ethical approval for this study was granted by the University of Zambia Biomedical Research Ethics Committee (UNZABREC; REF. No: 6453-2025) and the National Health Research Authority of Zambia (NHRA-2326/13/06/2025). Written informed consent was obtained from all participants prior to data collection. For participants younger than 18 years, written assent was obtained from the participant and written guardian consent was obtained from a parent or legal guardian. Participation was voluntary and withdrawal at any stage carried no consequences for access to healthcare services. The collected data was stored in such a way that it could not identify individuals, while ensuring confidentiality.

## Results

### Characteristics of women

The determined sample size for this study was 428 but only 413 were eligible and consented to participate in the survey due to time constraints and childcare responsibilities, giving a response rate of 96% (Figure: 1).

Most respondents were aged 20-24 years, followed by those aged 25-29 years, while adolescents aged 15-19 years represented the smallest proportion of the sample. The majority of participants were married, and most mothers reported having one or two children, indicating relatively low parity among the study population. Religious affiliation was predominantly Christian, reflecting the broader demographic profile of urban Zambia.

The sample comprised 27.4% adolescents (15–19 years), 33.2% aged 20-24 years, and 39.5% aged 25–29 years. Most participants were married (57.5%), followed by single (36.9%), separated (3.2%), and widowed (2.4%). Regarding parity, 20.0% were primiparous, 38.2% had one prior child, 31.1% had two, and 10.7% had three or more children. Religious affiliation was predominantly Christian (97.3%).

### Factors associated with EBF

The study population showed moderate educational attainment but substantial economic vulnerability, with most mothers unemployed and only a small proportion in formal employment. Despite this, nearly all participants reported receiving neonatal care education during antenatal or postnatal services. Cultural and psychosocial factors varied across respondents, including engagement with traditional practices, consultation with traditional healers, peer support participation, and postnatal mental health experiences, all of which were examined for their association with exclusive breastfeeding.

The study population demonstrated moderate educational attainment (50% secondary or higher) with 61.7% unemployed and nearly 80% earning K1,000-K5,000 monthly. Formal employment was uncommon (8.5%), suggesting substantial economic dependency and constrained financial autonomy. Despite these structural constraints, nearly universal exposure to neonatal education (99.5%) and awareness of recommended practices (98.3%) was observed.

Traditional influence persisted primarily through consultation rather than practice: while 78.5% consulted traditional healers or elders, 75.7% reported never using traditional neonatal practices, and only 9.1% indicated cultural restrictions that prevented recommended care. This divergence suggests that elder consultation may function more as social support than strict behavioural endorsement, though social desirability bias cannot be excluded.

Psychosocial vulnerability was notable, with 25.3% reporting frequent or very frequent postnatal anxiety, and 88.1% acknowledging that mental health influences neonatal care. Peer support was nearly universal (97.6%), limiting its variability as a differentiating factor.

Overall, the descriptive profile indicates a population with high informational exposure and strong social networks, yet operating within economic constraints and measurable psychosocial stress conditions that provide critical context for interpreting determinants of exclusive breastfeeding.

### Neonatal care practices

#### Frequency of EBF

The distribution reflects both knowledge of optimal feeding practices and the extent to which these practices are implemented in daily care routine. Nearly all mothers (98.1%) reported breastfeeding at the recommended frequency of every 2-3 hours, indicating high adherence to immediate feeding guidance. However, sustained adherence to EBF for the first six months was lower, with 71.6% reporting compliance and 28.4% not adhering to the recommendation (Figure 2: Distribution of knowledge on breastfeeding frequency among mothers).

**Fig 2:**
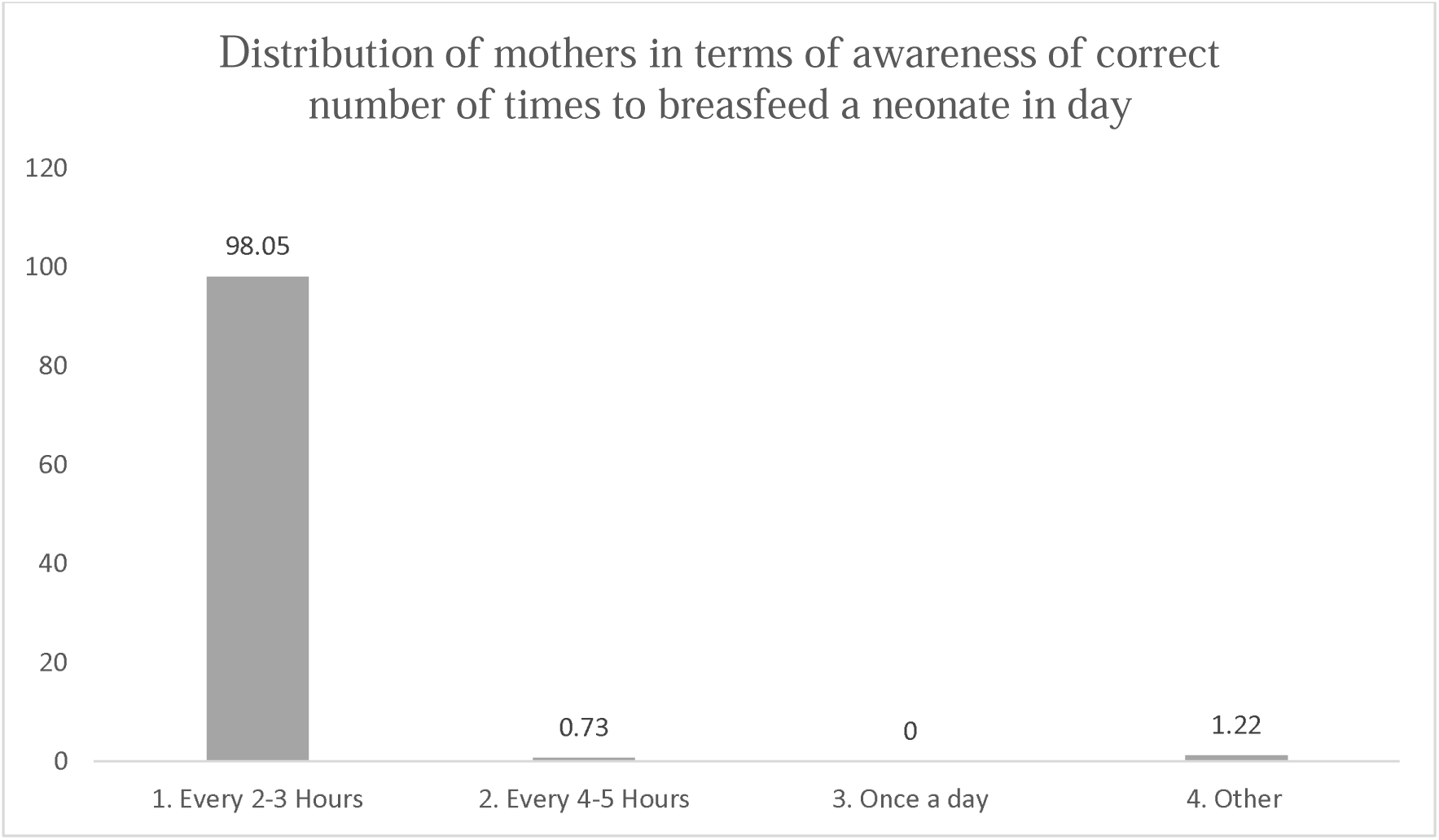
Aware of correct frequency of breastfeeding a neonate.

#### Prevalence of EBF

When interpreted alongside the near-universal awareness of exclusive breastfeeding (98.3%) and receipt of neonatal education (99.5%), the observed 71.6% EBF prevalence indicates an approximate 27-percentage-point knowledge-practice gap. This divergence suggests that although early breastfeeding behaviours are widely initiated, sustained exclusive breastfeeding is shaped by influences beyond knowledge exposure alone, underscoring the role of structural, cultural, and psychosocial determinants in neonatal care practices.

Although this prevalence exceeds the global average of approximately 48% among infants under six months, it does not necessarily translate into sustained adherence to the World Health Organization recommendation of exclusive breastfeeding for the full six-month period (Figure 3: Prevalence of exclusive breastfeeding).

**Fig 3:**
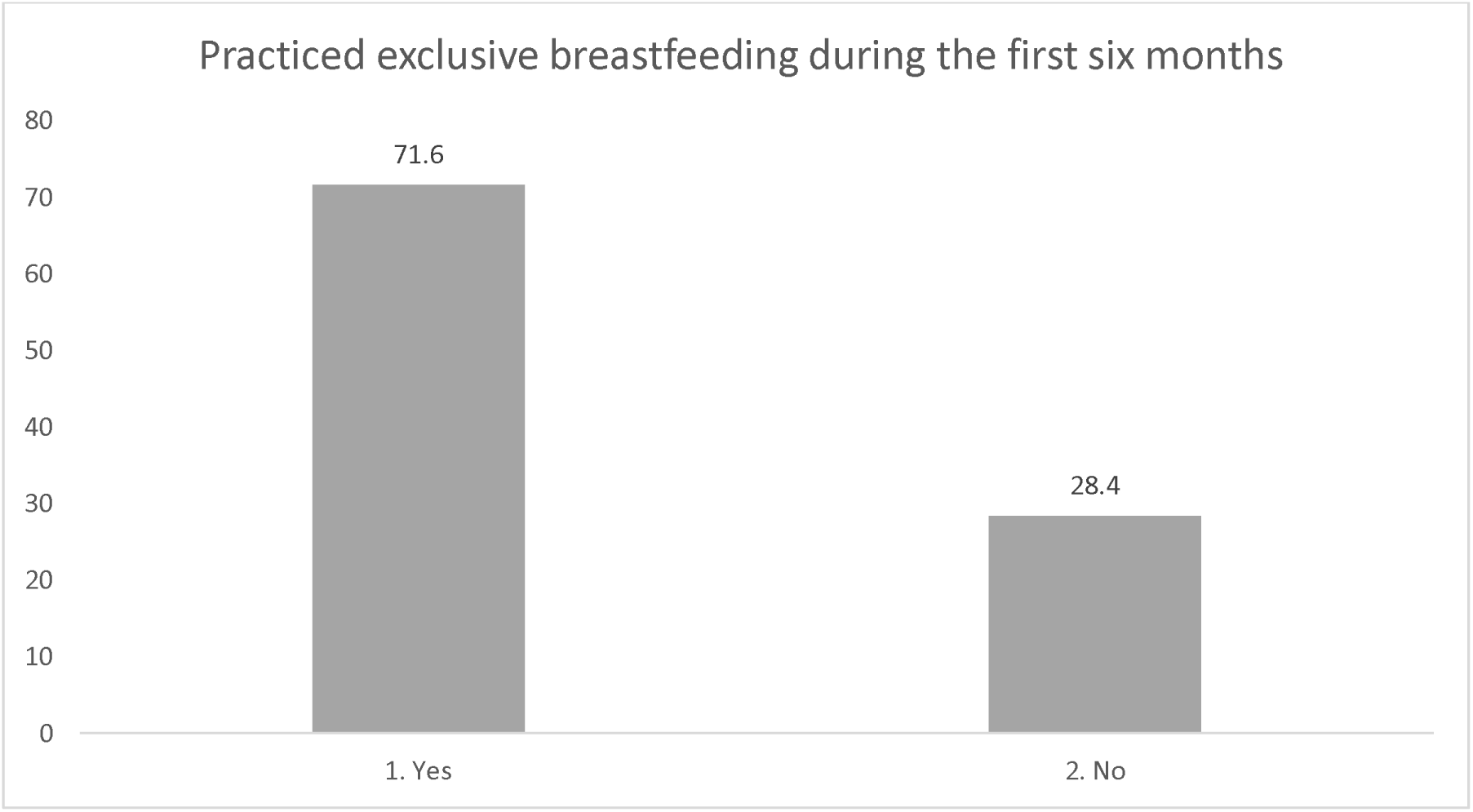
Distribution of mothers who practiced exclusive breastfeeding.

### Crude and adjusted logistic regression analysis

Multivariable analysis identified maternal age, marital status, employment, traditional practices, consultation with traditional healers, and postnatal anxiety as significant predictors of exclusive breastfeeding. Mothers aged 25-29 years had lower odds of EBF compared with adolescents, while married and formally employed mothers showed higher odds of practicing EBF.

Avoidance of traditional neonatal practices and traditional healer consultations was strongly associated with increased EBF likelihood. In contrast, postnatal anxiety significantly reduced the odds of exclusive breastfeeding, highlighting the importance of psychosocial factors in breastfeeding behaviour.

#### Model diagnostics

The Hosmer-Lemeshow goodness-of-fit test indicated adequate model fit (χ²(8)=5.18, p=0.738). No evidence of problematic multicollinearity was observed (VIF <10 for all variables).

In unadjusted analyses, several demographic, socioeconomic, cultural, and psychosocial factors were significantly associated with exclusive breastfeeding (EBF). Mothers aged 25-29 years had substantially lower odds of EBF compared to adolescents aged 15-19 years (OR=0.33, 95% CI: 0.18-0.59, p<0.001), and increasing parity showed a strong inverse association, with progressively lower odds among mothers with one, two, or three or more children (all p≤0.001).

Higher educational attainment was also inversely associated with EBF (all p≤0.002). In contrast, unemployment (OR=2.07, 95% CI: 1.30-3.29, p=0.002) and earning K1,000-K5,000 monthly (OR=2.09, 95% CI: 1.14-3.85, p=0.018) were positively associated with EBF.

Avoidance of traditional neonatal practices demonstrated strong associations, including not using traditional methods (OR=0.05, 95% CI: 0.02-0.11, p<0.001) and not consulting traditional healers (OR=0.06, 95% CI: 0.03-0.11, p<0.001).

Postnatal anxiety showed a pronounced graded inverse relationship with EBF across all frequency categories (all p<0.001). Awareness of neonatal care practices and receipt of neonatal education were not significantly associated with EBF.

After multivariable adjustment, independent determinants of EBF included younger maternal age, marital status, formal employment, avoidance of traditional practices, and lower levels of postnatal anxiety.

Mothers aged 25-29 years remained less likely to practice EBF compared to those aged 15-19 years (AOR=0.17, 95% CI: 0.03-0.99, p=0.048), while age 20-24 was not significant. Married (AOR=4.83, 95% CI: 1.59-14.65, p=0.005) and separated mothers (AOR=13.66, 95% CI: 1.89-98.71, p=0.010) had significantly higher odds of EBF compared to single mothers.

Formal employment was positively associated with EBF (AOR=3.94, 95% CI: 1.12-13.85, p=0.033). Cultural factors remained strong predictors: not using traditional neonatal practices (AOR=0.14, 95% CI: 0.04-0.53, p=0.004) and not consulting traditional healers (AOR=0.06, 95% CI: 0.02-0.18, p<0.001) were independently associated with EBF.

On the other hand, the crude odds ratio (OR) for secondary education relative to no formal education (OR = 0.01; 95% CI: 0.002–0.10) observed in the standard logistic regression is implausibly extreme and indicative of quasi-complete separation. This occurs when a predictor variable nearly perfectly predicts the outcome, resulting in unstable and biased maximum likelihood estimates. Such estimates are not substantively meaningful and may mislead interpretation if presented without adjustment.

To address this limitation, Firth’s penalised logistic regression was applied as a bias-reduction method. This approach corrects for small-sample bias and separation by introducing a penalty to the likelihood function, thereby producing finite and more reliable parameter estimates. Firth regression is particularly appropriate in settings with sparse data or rare outcome categories.

#### Comparison of Standard and Penalised Estimates

A comparison of the standard and Firth-adjusted crude models revealed substantial differences. In the standard model, education categories exhibited extremely small ORs with narrow confidence intervals (e.g., secondary education: OR = 0.0135; 95% CI: 0.0018-0.1003), suggesting an unrealistically strong protective effect. In contrast, the Firth-adjusted estimates were notably attenuated (secondary education: OR = 0.21; 95% CI: 0.03-1.64) and no longer statistically significant. Similar attenuation patterns were observed for primary and tertiary education.

#### Interpretation of Revised Estimates

The Firth-adjusted results indicate that the previously observed strong association between higher education and the outcome was likely driven by sparse data bias rather than a true effect. While the direction of association remains suggestive of a protective relationship, the wide confidence intervals and lack of statistical significance indicate greater uncertainty and reduced precision. This suggests that the evidence for an independent effect of education at the crude level is limited.

#### Implications for Analysis and Reporting

The penalised regression estimates were prioritised for variables affected by separation. Where appropriate, standard logistic regression results were supplemented with Firth estimates, and this approach has been clearly indicated in table 4. This ensures that reported findings are robust, interpretable, and not influenced by statistical artefacts.

**Table 1:**
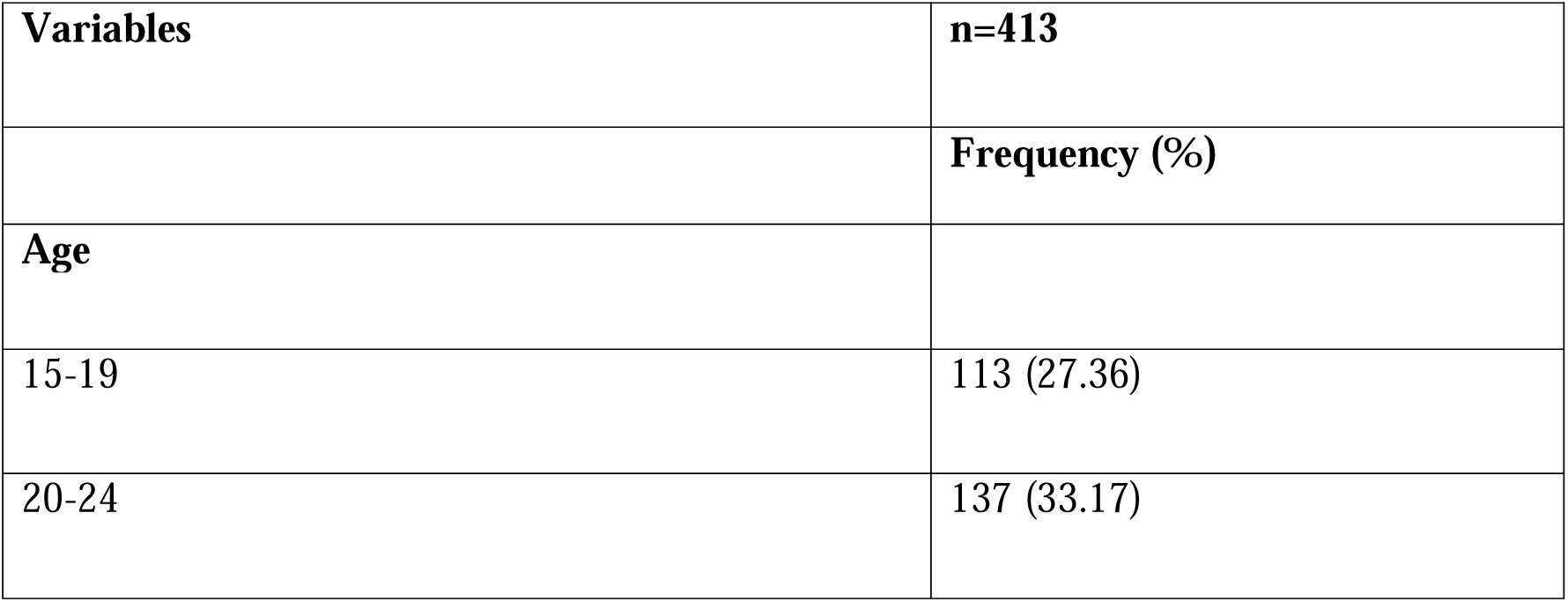
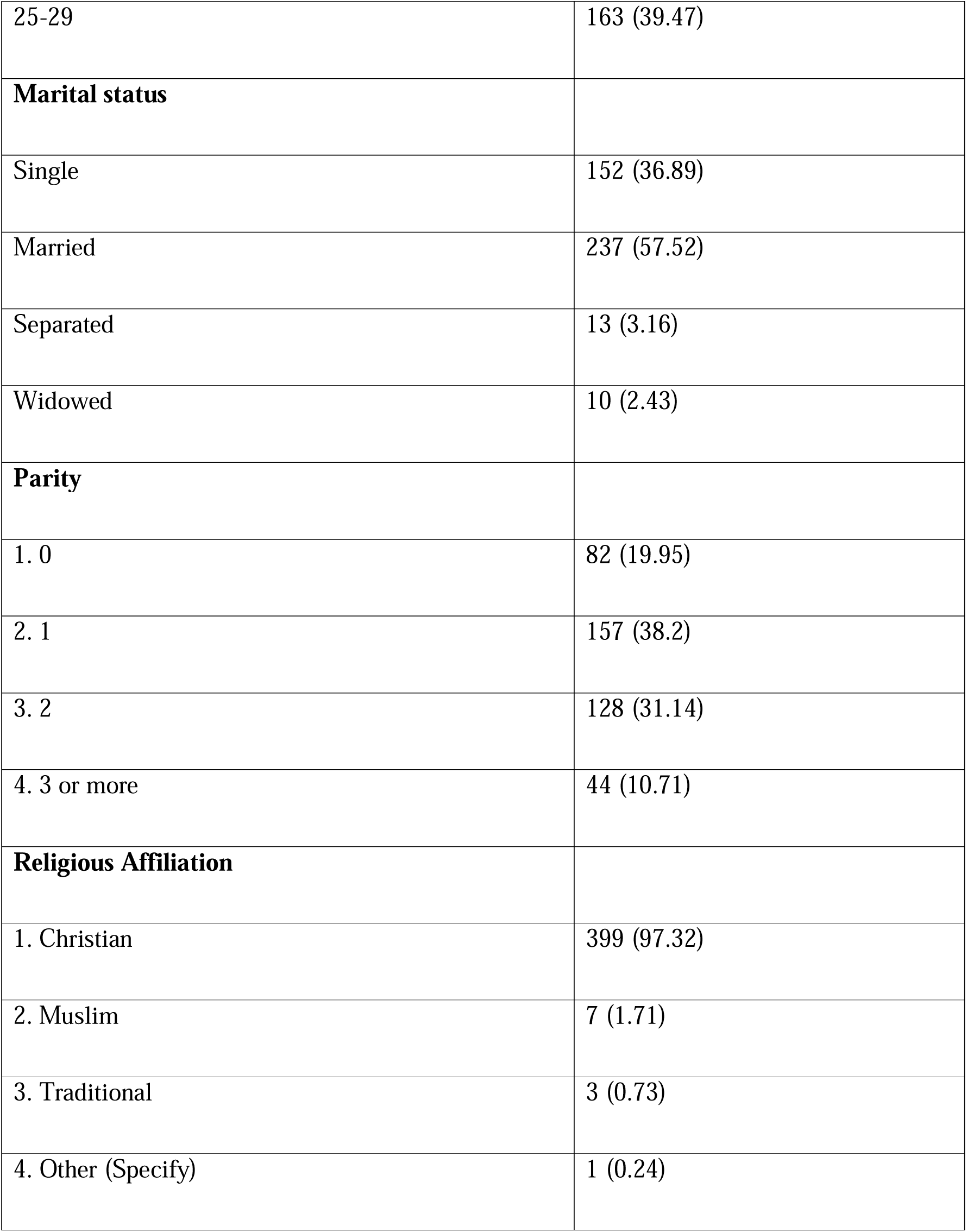
Demographic characteristics of women.

**Table 2:** Frequency distribution of factors associated with EBF.

| Domain | Variable | n =413) |
| --- | --- | --- |
|  |  | Frequency (%) |
| **Socioeconomic factors** | **Education** |  |
|  | No formal education | 84 (20.5) |
|  | Primary | 119 (29.0) |
|  | Secondary | 121 (29.5) |
|  | Tertiary | 86 (21.0) |
|  | **Occupation** |  |
|  | Informal employment | 123 (29.9) |
|  | Formal employment | 35 (8.5) |
|  | Unemployed | 254 (61.7) |
|  | **Monthly income** |  |
|  | < K1,000 | 51 (12.5) |
|  | K1,000–K5,000 | 327 (80.0) |
|  | K5,000–K10,000 | 31 (7.6) |
| **Traditional factors** | **Existence of traditional methods in the community** |  |
|  | Yes | 116 (28.2) |
|  | No | 296 (71.8) |
|  | **Use of specific traditional neonatal practices** |  |
|  | Yes | 163 (39.7) |
|  | No | 248 (60.3) |
|  | **Consult traditional healers/elders** |  |
|  | Yes | 322 (78.5) |
|  | No | 88 (21.5) |
|  | **Frequency of tradition practices** |  |
|  | Never | 312 (75.7) |
|  | Not often | 40 (9.7) |
|  | Often | 59 (14.3) |
|  | Very often | 1 (0.2) |
|  | **Traditional beliefs prevent recommended EBF** |  |
|  | Yes | 37 (9.1) |
|  | No | 372 (90.9) |
| **Psychosocial factors** | **Anxiety during PNC** |  |
|  | Never | 193 (47.0) |
|  | Not often | 114 (27.7) |
|  | Often | 84 (20.4) |
|  | Very often | 20 (4.9) |
|  | Mental health affects neonatal care |  |
|  | Yes | 362 (88.1) |
|  | No | 49 (11.9) |
| **Peer support** | **Received emotional support during PNC** |  |
|  | Yes | 402 (97.6) |
|  | No | 10 (2.4) |
| **Knowledge factors** | **Aware of neonatal care practices** |  |
|  | Yes | 404 (98.3) |
|  | No | 7 (1.7) |
|  | **Received neonatal care education** |  |
|  | Yes | 408 (99.5) |
|  | No | 2 (0.5) |

**Table 3:**
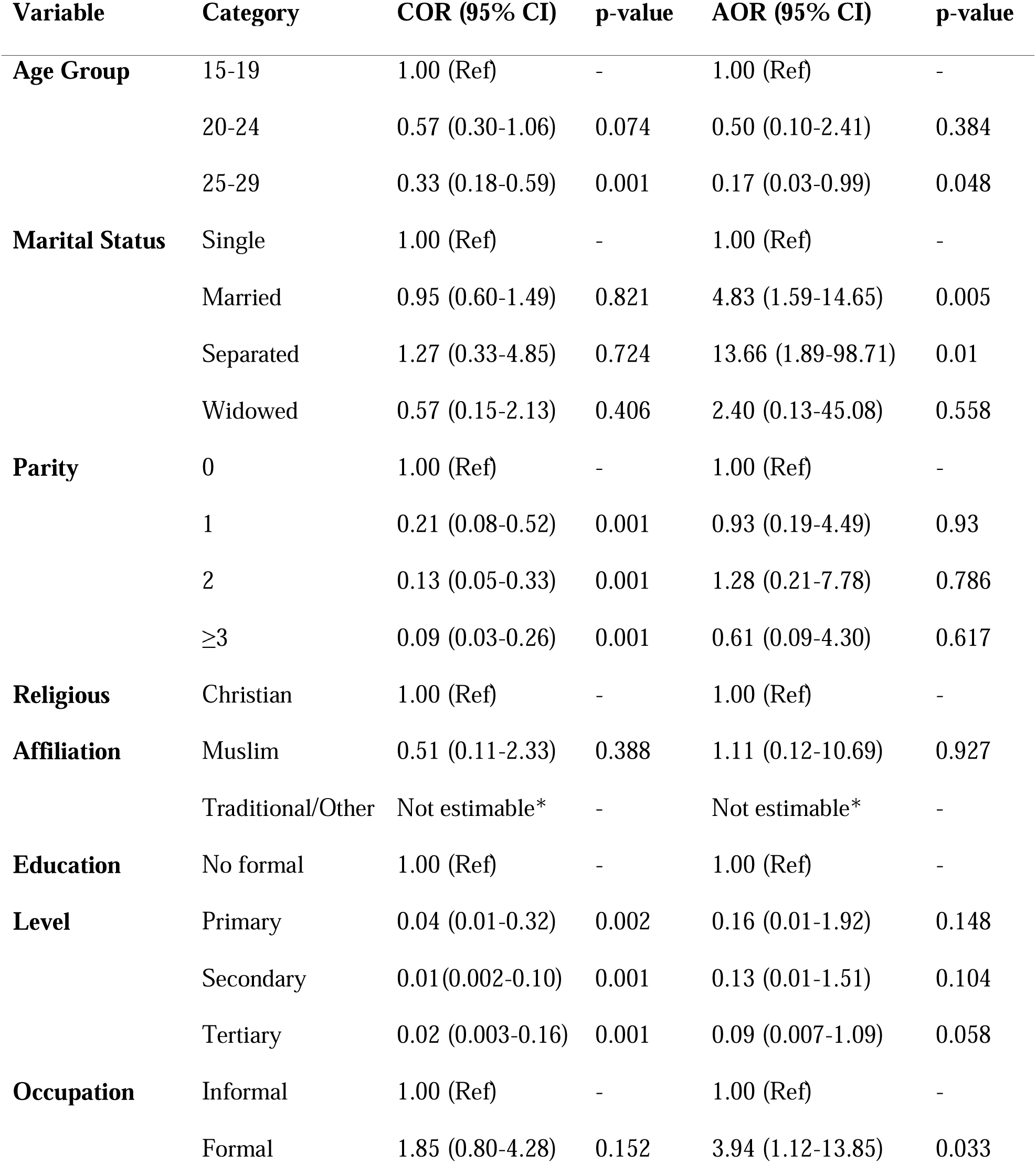
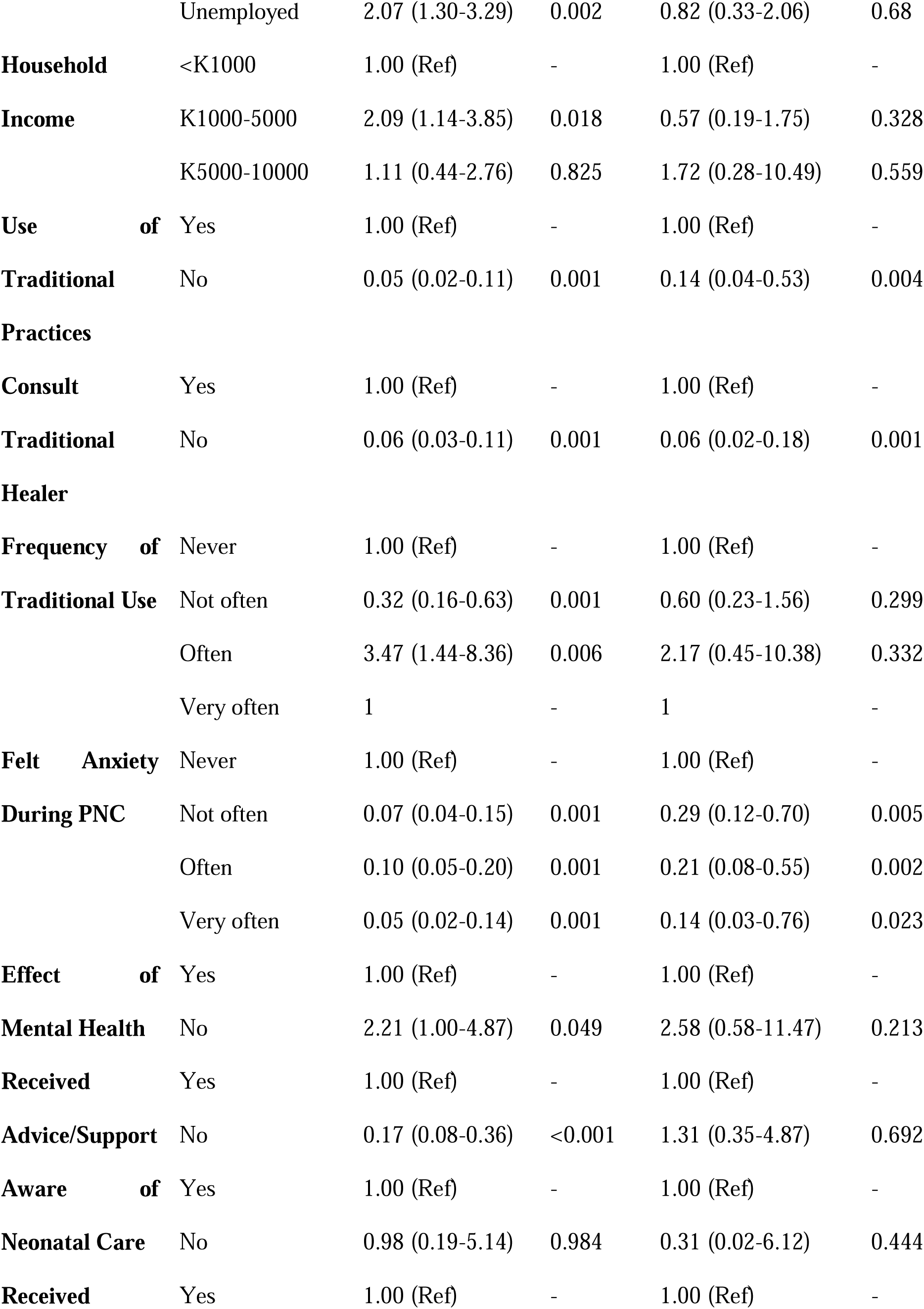
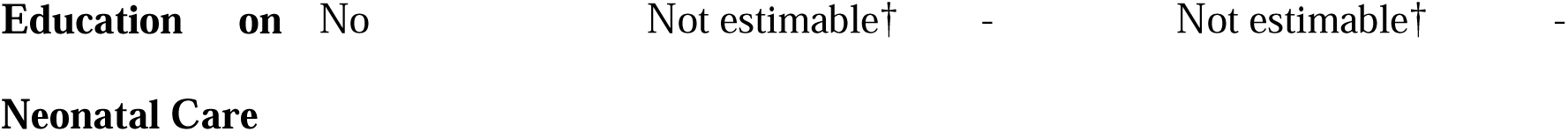
Crude and adjusted logistic regression analysis.

**Table 4:** Firth’s Penalised Logistic regression.

| Variables | Categories | COR (95% CI) | P. Value |
| --- | --- | --- | --- |
| **Age Group** | 15-19 | 1.00 (Ref) |  |
|  | 20-25 | 0.55 (0.13, 2.38) | 0.428 |
|  | 25-29 | 0.22 (0.04, 1.07) | 0.061 |
| **Marital Status** | Not Married | 1.00 (Ref) |  |
|  | Married | 3.94 (1.41, 11.00) | 0.009 |
|  | Separated | 9.47 (1.55, 57.88) | 0.015 |
|  | Widowed | 2.00 (0.13, 31.95) | 0.623 |
| **Parity** | Number_of_Children | 1.00 (Ref) |  |
|  | 1 | 0.90 (0.22, 3.74) | 0.888 |
|  | 2 | 1.23 (0.24, 6.39) | 0.802 |
|  | >3 | 0.65 (0.11, 3.90) | 0.635 |
| **Religious Affiliation** | Christian | 1.00 (Ref) |  |
|  | Muslim | 1.17 (0.16, 8.78) | 0.881 |
|  | Traditional religion | 0.03 (0.001, 1.40) | 0.074 |
|  | Other (specify) | 1.01 (0.03, 30.48) | 0.997 |
| **Education Level** | No formal education | 1.00 (Ref) |  |
|  | Primary education | 0.26 (0.03, 2.04) | 0.199 |
|  | Secondary education | 0.21 (0.03, 1.64) | 0.137 |
|  | Tertiary education | 0.15 (0.02, 1.27) | 0.082 |
| **Occupation** | Informal | 1.00 (Ref) |  |
|  | Formal employment | 3.36 (1.04, 10.88) | 0.043 |
|  | Unemployed | 0.87 (0.37, 2.02) | 0.725 |
| **Household Income** | <K1000 | 1.00 (Ref) |  |
|  | K1000-K5000 | 0.63 (0.22, 1.79) | 0.386 |
|  | K5000-K10000 | 1.56 (0.29, 8.37) | 0.604 |
| **Use of Traditional Practices** | Yes | 1.00 (Ref) |  |
|  | No | 0.10 (0.06, 0.66) | 0.009 |
| **Consult Traditional Healer** | Yes | 1.00 (Ref) |  |
|  | No | 0.09 (0.04, 0.25) | 0 |
| **Frequency of Traditional use** | Never | 1.00 (Ref) |  |
|  | Not often | 0.63 (0.26, 1.53) | 0.305 |
|  | Often | 1.77 (0.43, 7.21) | 0.428 |
|  | Very often | 0.02 (0.0005, 1.25) | 0.065 |
| **Felt Anxiety During PNC** | Never | 1.00 (Ref) |  |
|  | Not often | 0.33 (0.13, 0.76) | 0.008 |
|  | often | 0.25 (0.10, 0.62) | 0.003 |
|  | Very often | 0.18 (0.04, 0.83) | 0.028 |
| **Effect of Mental Health** | Yes | 1.00 (Ref) |  |
|  | No | 2.15 (0.54, 8.51) | 0.274 |
| **2.Received Advice or Support** | Yes | 1.00 (Ref) |  |
|  | No | 1.25 (0.37, 4.20) | 0.718 |
| **Aware of Neonatal Care Practices** | Yes | 1.00 (Ref) |  |
|  | No | 0.39 (0.03, 5.15) | 0.478 |
| **Received Education on Neonatal Care Practices** | Yes | 1.00 (Ref) |  |
|  | No | 0.69 (0.01, 68.71) | 0.874 |

#### Assessment of interaction between postnatal anxiety and marital status

PNC anxiety and marital status emerged as strong independent predictors and this raised concerns as to whether this was due to interaction between the two variables. To evaluate potential effect modification, a multiplicative interaction between postnatal anxiety and marital status was tested. Given evidence of sparse data and separation, the interaction model was estimated using Firth’s penalised logistic regression.

The results from the joint test for interaction were not statistically significant (χ² = 12.29, p = 0.198), indicating no evidence that marital status modifies the association between postnatal anxiety and exclusive breastfeeding. Although some interaction coefficients were large, they were imprecise with wide confidence intervals, reflecting small subgroup sizes.

Conditional marginal effects were plotted to visually assess the interaction (Figure 4). The results show that predicted effects of postnatal anxiety on exclusive breastfeeding are broadly similar across marital status categories. While minor variations are observed, the confidence intervals overlap substantially, indicating no meaningful differences between groups.

**Fig 4:**
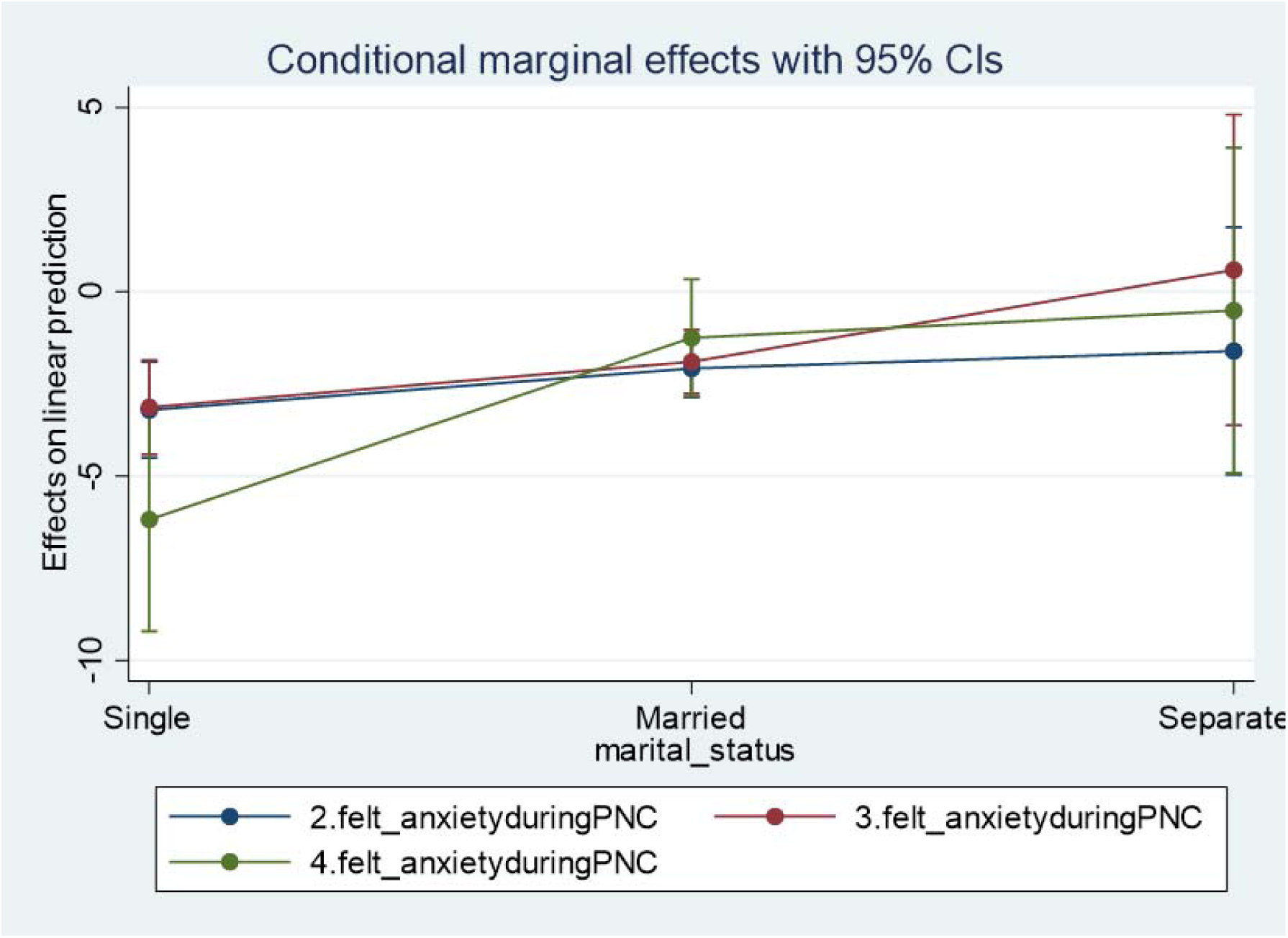
Conditional marginal effects with 95% CIs.

Across all marital categories, increasing levels of postnatal anxiety were associated with reduced predicted likelihood of exclusive breastfeeding. Wider confidence intervals in some groups, particularly among separated mothers, reflect limited sample sizes rather than true heterogeneity of effects. These findings are consistent with the regression analysis and support the absence of interaction.

Overall, the findings indicate that postnatal anxiety is a key determinant of exclusive breastfeeding practices, while marital status does not significantly influence this relationship. The use of Firth regression strengthened the validity of these findings by correcting for sparse data bias and ensuring stable estimation.

#### Summary

Postnatal anxiety retained a graded inverse association (AOR range: 0.14-0.29) while education, income, parity, peer support, and neonatal care knowledge were not independently associated with EBF after adjustment. Some crude odds ratios, particularly for maternal education (e.g., secondary versus no formal education, OR = 0.01), were extremely small and likely reflect sparse cell counts and quasi-complete separation in these subgroups.

Similarly, a few variables were not estimable due to insufficient observations in certain categories. However, a sensitivity analysis using penalized logistic regression (Firth’s method) was performed to address sparse-data bias and improve model stability. The application of Firth penalised regression strengthens the validity of the analysis by addressing bias arising from sparse data. This approach enhances the reliability of the findings and supports more accurate interpretation of associations within the study population

## Discussion

This study demonstrates that EBF (71.6%), alongside near-universal neonatal education exposure (99.5%) alone does not guarantee behavioural adherence among young mothers in peri-urban Lusaka. It is shaped more by psychosocial and sociocultural dynamics, a finding that challenges the predominance of knowledge-centric breastfeeding promotion strategies in this setting.

Multivariable analysis revealed that EBF was independently associated with age, marital status, formal employment, engagement in traditional neonatal practices, and postnatal anxiety, whereas maternal education, income, peer support, and knowledge were not predictive after adjustment. These findings highlight a persistent knowledge-practice gap and suggest that structural, sociocultural, and psychosocial factors play a more decisive role in shaping sustained breastfeeding behaviour than informational exposure alone.

### Demographic and social-economic factors

Mothers aged 25-29 years had significantly lower odds of EBF compared to younger mothers (AOR=0.17, p=0.048). While previous regional studies suggest older maternal age predicts improved neonatal care adherence^12,13^, our findings indicate that in peri-urban Lusaka, older young mothers may face increased economic and caregiving pressures that compete with sustained EBF. This divergence underscores the contextual variability of age effects and cautions against assuming uniform demographic gradients.

Marital status demonstrated a strong and independent association with EBF. Married (AOR=4.83, p=0.005) mothers were significantly more likely to practice EBF compared to single mothers. These results are consistent with evidence linking partner support to improved infant feeding outcomes^14,15^. The magnitude of association suggests that relational stability may provide both emotional reinforcement and practical support conducive to sustained breastfeeding.

Formal employment was positively associated with EBF (AOR=3.94, p=0.033), contrasting with earlier literature suggesting employment constrains breastfeeding due to workplace inflexibility^16^. In this setting, formal employment may proxy socioeconomic stability and access to maternity protections. Zambia’s labour regulations provide for maternity leave provisions in formal employment sectors^17^; however, enforcement and coverage may vary. The observed positive association between formal employment and exclusive breastfeeding may reflect access to structured maternity protections or predictable work environments compared to informal employment contexts.

Although education and income were not independently associated with EBF, unadjusted trends indicated lower odds among more educated mothers. This contrasts with regional findings linking education to improved newborn care practices^12,13^ and may reflect peri-urban dynamics where education coincides with workforce participation and time constraints.

### Traditional practices

Lack of ngagement in traditional neonatal practices was strongly and inversely associated with EBF (AOR=0.14, p=0.004; AOR=0.06, p<0.001). The magnitude of effect suggests substantial behavioural divergence between mothers adhering to biomedical guidance and those influenced by customary norms. Similar conflicts between traditional beliefs and exclusive breastfeeding recommendations have been documented in southern Africa^18,19^ and other sub-Saharan contexts where elder advice supersedes clinical counselling^20,21^.

These findings reinforce the need to situate breastfeeding promotion within sociocultural systems rather than Rather than positioning traditional neonatal practices as inherently oppositional to biomedical recommendations. Therefore, culturally responsive counselling approaches may offer opportunities for integration and respectful dialogue. However, given the cross-sectional design, it remains unclear whether traditional beliefs preceded or co-evolved with feeding practices.

### Maternal mental health

A graded association was observed between postnatal anxiety and reduced odds of EBF (AOR=0.29 for infrequent anxiety; AOR=0.14 for very frequent anxiety), suggesting a dose-response relationship. This aligns with literature demonstrating that maternal psychological distress impairs practice, while some studies in Sub-Saharan Africa indicated that it had no effect on EBF^22,23^. Anxiety may operate through diminished self-efficacy, perceived insufficient milk supply, cognitive overload, and reduced help-seeking behaviour.

In Zambia like other Low Middle Income Countries (LMIC), perinatal mental health remains under-integrated into primary maternal care^24^. Our findings provide empirical support for embedding brief mental health screening within postnatal services. Nevertheless, temporality cannot be established; anxiety may both influence and result from breastfeeding challenges.

### Peer support

The study did not find a statistically significant association between peer support and exclusive breastfeeding. However, peer support was reported by 97.6% of participants, resulting in minimal variability and a likely ceiling effect. Under such conditions, logistic regression has limited discriminatory power, and estimates may be biased toward the null due to insufficient contrast between exposure groups.

This limitation suggests that the null finding should be interpreted with caution and does not necessarily indicate the absence of a meaningful relationship. It is plausible that peer support is broadly available in this setting but varies in quality, intensity, or timing, dimensions not captured by the binary measure used in this study.

Future research should employ more granular measures of peer support (e.g., frequency, source, perceived usefulness) and ensure adequate variability to better assess its role in influencing neonatal care practices.

### Knowledge-practice gap

The prevalence of exclusive breastfeeding (EBF) in this study (71.6%) was slightly higher than the national estimate of approximately 64% among infants aged 0-5 months reported in the ZDHS 2024. This difference may partly reflect the facility-based sampling frame, as mothers attending postnatal services are more likely to receive breastfeeding counselling and support than the general population. Nonetheless, the prevalence is broadly consistent with findings from urban and peri-urban settings in Zambia, where access to maternal and child health services and breastfeeding promotion programmes tends to be greater than in rural areas.

Despite near-universal exposure to neonatal care education, EBF practice remained lower (71.6%), indicating a 27-percentage-point implementation gap, and knowledge variables were not independently associated with EBF, consistent with evidence from other sub-Saharan African settings documenting knowledge-practice discordance^25,26^. This suggests that informational exposure alone is insufficient to ensure behavioural adoption. Mothers may recall recommendations but lack the practical capacity to address challenges such as perceived low milk supply or social pressures^27^, while structural constraints including income-generating demands and limited breastfeeding-supportive environments may further weaken the influence of knowledge on practice^28^.

These findings align with critiques of purely cognitive behaviour models, which may underestimate the role of social norms and psychosocial barriers in shaping maternal health behaviours^29^. Although the observed prevalence exceeds the global average of approximately 48% among infants under six months, it does not guarantee sustained adherence to the World Health Organization recommendation of exclusive breastfeeding for the first six months of life.

Overall, the results indicate that EBF in this peri-urban population is influenced by a complex interaction of social, cultural, and psychosocial factors, including maternal age, marital status, traditional practices, and postnatal anxiety. Improving breastfeeding outcomes therefore requires approaches that extend beyond information provision to address psychosocial wellbeing and culturally embedded practices, including the integration of mental health screening, culturally sensitive counselling, and family-centred breastfeeding support within routine postnatal services.

### Implications

The findings indicate that improving EBF in peri-urban Zambia requires integrated strategies addressing psychosocial and sociocultural barriers in addition to health education. Interventions should incorporate mental health screening, engage partners and elder family members, strengthen behaviour change communication approaches that build practical skills, and advocate for breastfeeding-supportive employment environments. Peer-support models implemented in East Africa demonstrate potential scalability^30,31^.

### Limitations

This study has several limitations. The cross-sectional design precludes causal inference and does not allow definitive determination of temporal sequencing; reverse causality cannot be excluded. It is plausible that breastfeeding difficulties may contribute to heightened anxiety symptoms, just as anxiety may impair exclusive breastfeeding adherence. Longitudinal studies are therefore warranted to clarify the directionality of this association. Facility-based sampling limits generalisability to mothers not accessing postnatal services, and the sampling interval of one approximated near-consecutive recruitment at one facility, introducing elements of convenience sampling that may affect representativeness. Self-reported breastfeeding practices may be subject to recall and social desirability bias, and maternal anxiety was measured using a self-report scale rather than a diagnostic instrument, which may affect measurement precision. Additionally, some subgroup estimates yielded wide confidence intervals and should be interpreted cautiously due to limited statistical stability. The limited variability in peer support exposure constrained its usefulness as a predictor in regression models, potentially masking true associations. Nevertheless, peri-urban communities represent rapidly expanding demographic contexts in sub-Saharan Africa, suggesting that the findings retain relevance for similar transitional urban environments.

## Conclusion

In this peri-urban Zambian setting, psychosocial and cultural determinants rather than knowledge deficits emerge as primary drivers of EBF practice among high-risk young mothers. EBF adherence was independently associated with marital support, formal employment, avoidance of traditional neonatal practices, and maternal mental health status, while knowledge alone did not predict behaviour. Notably, postnatal anxiety demonstrated a strong and graded negative association with EBF, indicating that psychosocial vulnerability may substantially undermine optimal infant feeding practices. Collectively, these findings suggest that EBF practice is shaped less by informational deficits than by relational, cultural, structural, and psychological determinants. Breastfeeding promotion strategies in similar peri-urban contexts should therefore move beyond information-centric approaches toward integrated models that incorporate culturally responsive counselling, partner engagement, structural support mechanisms, and routine maternal mental health screening within postnatal services. Further longitudinal cohort studies are needed to establish the temporal direction of the anxiety-EBF relationship and to evaluate the effectiveness of integrated psychosocial and culturally responsive postnatal interventions on EBF continuation rates.

## Data Availability

All data produced in the present study are available upon reasonable request to the authors

## Data availability

The dataset supporting the conclusions of this article contains sensitive participant information and is not publicly available. Anonymised data may be made available upon reasonable request to the corresponding author and with approval from the UNZABREC.

## Acknowledgments

The author acknowledges the participating mothers and health facility staff in Mandevu Constituency for their cooperation and support. Appreciation is extended to academic supervisors and colleagues who provided guidance during the study.

## Author Contributions

Conceptualization: Godwill Silupya, Kalonga Mwiinga, Rosemary N. Likwa

Methodology: Godwill Silupya, Kalonga Mwiinga

Formal analysis: Godwill Silupya

Investigation: Godwill Silupya

Data curation: Godwill Silupya

Writing-original draft: Godwill Silupya

Writing-review & editing: Rosemary N. Likwa, Kalonga Mwiinga, Twaambo Nkwendenda

Supervision: Rosemary N. Likwa, Kalonga Mwiinga, Twaambo Nkwendenda

Validation: Rosemary N. Likwa, Kalonga Mwiinga

Project administration: Godwill Silupya

## Notes

### Competing Interest Statement

The authors have declared no competing interest.

### Author Declarations

University of Zambia Biomedical Research Ethics Committee of University of Zambia gave ethical approval for this work. Ethics committee/IRB of National Health Research Authority waived ethical approval for this work

## References

1. World Health Organization (2020). Newborns: *Improving survival and well-being*. Geneva: WHO.

2. World Health Organization (2023). *WHO recommendations on maternal and newborn care for a positive postnatal experience*. Geneva: WHO.

3. Bhutta ZA, Das JK, Bahl R, Lawn JE, et al; Lancet Newborn Interventions Review Group; Lancet Every Newborn Study Group. Can available interventions end preventable deaths in mothers, newborn babies, and stillbirths, and at what cost? Lancet. 2014 Jul 26;384(9940):347–70. a. doi: 10.1016/S0140-6736(14)60792-3.

4. Lawn JE, Blencowe H, Oza S, You D, Lee AC, Waiswa P, Lalli M, Bhutta Z, Barros AJ, Christian P, Mathers C, Cousens SN; Lancet Every Newborn Study Group. Every Newborn: progress, priorities, and potential beyond survival. Lancet. 2014 Jul 12;384(9938):189–205. a. doi: 10.1016/S0140-6736(14)60496-7.

5. Zambia Statistics Agency (ZamStats), Ministry of Health (MOH) Zambia, and ICF. *Zambia Demographic and Health Survey* 2024. Lusaka, Zambia, and Rockville, Maryland, USA: ZamStats, MOH, and ICF; 2024.

6. Zambia Statistics Agency, Ministry of Health (MOH) Zambia, and ICF. (2018). *Zambia Demographic and Health Survey*. Lusaka, Zambia, and Rockville, Maryland, USA: Zambia Statistics Agency, Ministry of Health, and ICF.

7. Zambia Statistics Agency (2024). 2022 *Living Conditions Monitoring Survey Report*. Lusaka: Zambia Statistics Agency.

8. Jennifer L. Kelsey, Alice S. Whittemore, Alfred S. Evans, W. Douglas Thompson. Methods in Observation Epidemiology. Second Edition. Oxford University Press. 1996.

9. Greiner, T. Exclusive breastfeeding: measurement and indicators. Int Breastfeed J 9, 18 (2014). 10.1186/1746-4358-9-18

10. Clara Aarts, Elisabeth Kylberg, Agneta Hörnell, Yngve Hofvander, Mehari Gebre-Medhin, Ted Greiner, How exclusive is exclusive breastfeeding? A comparison of data since birth with current status data, *International Journal of Epidemiology*, Volume 29, Issue 6, December 2000, Pages 1041–1046, 10.1093/ije/29.6.1041

11. World Health Organization (WHO). (2008). Indicators for assessing infant and young child feeding practices: Part 1 Definitions. WHO. https://dev.nutritioncluster.net/sites/nutritioncluster.com/files/2020-01/Final_IYCF_prog_guide_May_26_2011.pdf

12. Thomas Chirwa, Chitalu Miriam, Chama-Chiliba. Essential newborn care practices in Zambia. Journal of Public Health in Africa. 10.4081/jphia.2022.2078

13. Chika Chioma Harriet Odira, Blessing Tochukwu Onyeje, Edith Anulika Udeogalanya, et al. Predictors of mothers’ home cord care, breastfeeding, and thermoregulation practices for newborns in a South-Eastern State, Nigeria. BioMed Centre Pregnancy and Childbirth vol: 25 issue: 1 year: 2025 doi: 10.1186/s12884-025-07705-x

14. Prost, A., et al. (2013). Women’s groups practicing participatory learning and action to improve maternal and newborn health in low-resource settings: A systematic review and meta-analysis. The Lancet, 381(9879), 1736–1746. 10.1016/S0140-6736(13)60685-6

15. Manda-Taylor, L., Mambulu, F., Malamba, F., Kunyenge, B., Phiri, T., Mwansambo, C., Kazembe, P., & Rosato, M. (2021). Exploring community support on safe motherhood: A case of Mchinji district, Malawi. African Journal of Primary Health Care and Family Medicine, 13(1), a2907. a. doi:10.4102/phcfm.v13i1.2907

16. Senarath, U. et al. (2007). Factors associated with maternal knowledge of new-born care among hospital-delivered mothers in Sri Lanka. Transactions of the Royal Society of Tropical Medicine and Hygiene, 101(8), pp. 823–830. a. 10.1016/j.trstmh.2007.03.003.

17. Employment Code Act No. 3 of 2019. Lusaka: Government of Zambia; 2019.

18. Buser, J. (2019). Cultural Practices, Knowledge, and Beliefs of Newborn Care and HealthSeeking in Rural Zambia. Thesis. http://deepblue.lib.umich.edu/handle/2027.42/151532.

19. Mwale, D., & Sitali, C. (2022). Peer support groups and their influence on maternal knowledge and practice of essential newborn care in Lusaka, Zambia. University of Zambia, 15(1), 1–10.

20. Tavares, E.A. de O. and Ramos, M.N. (2023). Cultural influence on Angolan maternal care of newborns and health strategies: health professionals’ perspective. Research, Society and Development, 12(4), pp. e9612441039–e9612441039. 10.33448/rsd-v12i4.41039.

21. Nankunda, J., Tumwine, J. K., & Tylleskär, T. (2021). The dual role of social support in promoting and hindering essential newborn care practices in Uganda. Journal of Health, Population and Nutrition, 40(1), 1–11.

22. Woldeyohannes, D., Tekalegn, Y., Sahiledengle, B., et al. (2021). Effect of postpartum depression on exclusive breastfeeding practices in Sub-Saharan Africa: A systematic review and meta-analysis. BioMed Central Pregnancy and Childbirth, 21, 113. a. 10.1186/s12884-020-03535-1

23. Neupane, S., Vuong, A., Haboush-Deloye, A., et al. (2025). Association between postpartum anxiety and depression and exclusive and continued breastfeeding practices. International Breasfeeding Journal, 20(1), 39. a. 10.1186/s13006-025-00734-4

24. Atif N, Lovell K, Rahman A. Maternal mental health: The missing “m” in the global maternal and child health agenda. Semin Perinatol. 2015;39(5):345–352. a. doi: 10.1053/j.semperi.2015.06.007.

25. Bee M, Shiroor A, Hill Z. Neonatal care practices in sub-Saharan Africa: a systematic review of quantitative and qualitative data. J Health Popul Nutr. 2018 Apr 16;37(1):9. a. doi: 10.1186/s41043-018-0141-5.

26. Sacks E, Moss WJ, Winch PJ, Thuma P, van Dijk JH, Mullany LC. Skin, thermal and umbilical cord care practices for neonates in southern, rural Zambia: a qualitative study. BioMed Central Pregnancy Childbirth. 2015 Jul 16;15:149. a. doi: 10.1186/s12884-015-0584-2.

27. Malinga S, Ilukena M, Chirwa T, Chama-Chiliba CM. Essential newborn care practices in Zambia. J Public Health Afr. 2022 Jul 26;13(2):2078. a. doi: 10.4081/jphia.2022.2078.

28. Phiri, T., Sitali, L., & Zulu, J. (2020). *Access to healthcare services and its impact on maternal knowledge of essential infant care practices in peri-urban Zambia*. University of Zambia Institutional Repository, 24(2), 63–72.

29. Janz NK, Becker MH. *The Health Belief Model*: a decade later. Health Educ Q.;11(1):1–47. 1984.

30. Eluri, S., Baliga, B.S., Rao, S.S. et al. Can Flip-Chart Assisted Maternal Education Improve Essential New Born Care Knowledge and Skills? A Randomized Controlled Trial. Matern Child Health J 26, 1891–1906 (2022). 10.1007/s10995-022-03409-2

31. Wright, J.L. et al. (2023). Design and implementation of a community-based mother-to-mother peer support programme for the follow-up of low birthweight infants in rural western Kenya. Frontiers in Paediatrics, p11. 10.3389/fped.2023.1173238.

